# Diagnosis of T-Cell-Mediated Kidney Rejection by Biopsy-Based Proteomics and Machine Learning

**DOI:** 10.1101/2020.05.11.20098285

**Authors:** Fei Fang, Peng Liu, Yang Zhao, Rajil Mehta, George Tseng, Parmjeet Randhawa, Kunhong Xiao

## Abstract

**Purpose:** This study is aimed at developing a clinic-friendly proteomics protocol and a machine learning (ML)-based molecular diagnostic test for T-cell-mediated rejection (TCMR) using formalin-fixed, paraffin-embedded (FFPE) biopsies.

**Experimental design:** Based on the procedures we reported for proteomic profiling of FFPE biopsies using Tandem Mass Tag (TMT)-based technology, a label-free-based quantitative proteomics protocol was developed as a more clinical-practical and cost-efficient molecular diagnostic test for renal transplant injection. This new protocol was applied to a set of FFPE biopsies from renal allograft injury patients and normal controls, including 5 TCMR, 5 polyomavirus BK nephropathy (BKPyVN) and 5 stable graft function (STA). Three different machine learning algorithms, linear discriminant analysis (LDA), support vector machine (SVM) and random forests (RF), were tested to build a prediction model for TCMR.

**Results:** About 750-1250 proteins were identified and quantified in each sample with high confidence using the label-free-based proteomics protocol. 178, 450 and 281 proteins were defined as differential expression (DE) proteins for TCMR vs STA, BKPyVN vs STA and TCMR vs BKPyVN, respectively. By comparing the quantitative data from the TMT- and label-free-based proteomics profiling, a classifier panel comprised of 234 DE proteins commonly quantified by two methods was generated to test different ML algorithms. Leave-one-out cross-validation result suggested that the RF-based model achieved the best prediction power for TCMR at both proteome and transcriptome level.

**Conclusions and clinical relevance:** Proteomics profiling of FFPE biopsies using a platform integrated of label-free quantitative proteomics with ML-based predictive model can help to discover biomarker panels and provide clinical molecular diagnostic tests to enhance biopsy interpretation for renal allograft rejection.

**Clinical Relevance:** This study is to develop a molecular diagnostic test for kidney rejection. An easy-to-use and cost-efficient protocol using label-free quantitative strategy was developed to profile proteome of FFPE biopsies from kidney allografts. A list of 234 DE identified from TCMR, BKPyVN and STA was generated as a classifier panel for these different phenotypes. This classifier panel was subjected to the optimized ML model, achieving high accuracy among both positive and negative control. This proof-of-principle study demonstrated the clinical feasibility of implementation of molecular diagnostic tests integrated of label-free-based quantitative proteomics and ML-derived disease predictive models to enhance biopsy interpretation for kidney transplantation patients. More accurate and specific molecular tests can lead to more effective treatment, prolong graft life, and improve the quality of life for patients with chronic kidney failure.

## 1. INTRODUCTION

In the United States alone, over 200,000 people are now living with functioning kidney transplants and rejection is the major cause for transplant loss [1]. There is an urgent need to evaluate the changes in rejection risks over time for T-cell-mediated rejection (TCMR), an important event in organ transplantation and a classic model for T-cell-mediated inflammatory diseases. With contemporary immunosuppression, TCMR is less frequent but remains the dominant early rejection phenotype and the end point in many clinical trials [2]. At present, this disease is mainly diagnosed with Banff lesion score *i* (Interstitial inflammation) to evaluate the degree of inflammation in nonscarred areas of cortex, which is a subjective and non-quantitative interpretation that requires experienced pathologists [3, 4], with significant inter-observer variability in multicenter clinical trials for diagnosis of TCMR.

Finding disease diagnostic patterns with “predictive power” is of great clinical value to enhance biopsy interpretation and to identify patients who may most likely benefit from a given treatment. In comparison with other biological materials, the formalin-fixed paraffin-embedded (FFPE) specimen has its unique traits in clinical diagnostics because of technical ease and low storage cost [5]. It was reported that gene expression profiling, using DNA- and RNA-level markers sourced from FFPE blocks, can be used as tools to diagnose and differentiate various cancers [6, 7]. However, attempts to implement these DNA- and RNA-based tests on a large scale in clinical setting have brought the realization that in practice these tests have their limitations. One limitation is that these technologies become dependent analysis of a small tissue fragment taken from a longer core sent for routine histology, with which easily missed the core with real disease information, as exemplified the observation that if two separate biopsy cores are taken from patients with BKV nephropathy (BKPyVN), viral inclusions can be seen in only one core in ~40% of samples [8]. Another important factor that limited the wide application of DNA- and RNA-based tests is their high cost in clinical practice.

As another important usage of FFPE blocks, proteomic profiling has been postulated as a "molecular microscope" to give better insight into the classification of renal transplant pathology [9]. Comparing to traditional diagnostic methods, proteomics-based tests has many advantages including superior specificity, sensitivity, and accuracy; high-throughput; capability of simultaneously monitoring multiple biomarkers, as well as low cost. Therefore, there is a high potential by developing biopsy-based proteomics tests to monitor kidney transplants and predict renal allograft injuries. However, there are few proteome studies on differentiating renal transplant disease phenotypes with FFPE biopsies due to the challenge in sample preparation to the small amount of formalin-induced cross-linking of proteins and screening out the real biomarkers from hundreds to thousands of differential expressed proteins quantified with the traditional quantitative proteomics.

In our latest work, the proteins were efficiently extracted from the FFPE biopsies by a combination of sequential mechanical mincing followed by sonication and heating. In combination with a Tandem Mass Tag (TMT)-based labeling protocol, a quantitative proteomic platform was successfully developed for proteomic profiling of FFPE biopsies [10]. Since the TMT-based quantitative proteomics protocol requires labeling of biopsy peptides with expensive isobaric TMT reagents and follow-up peptide fractionation, a proteomic profiling assay using this TMT-based protocol in the “real world” clinical setting will have its limitation. In this work, a more cost-efficient and technical practical protocol using label-free-based proteomics profiling technology integrated with efficient protein extraction strategy was developed to obtain useful protein panel for subsequent prediction.

Supervised machine learning (ML) algorithms have been a dominant method in the disease prediction field since it is well suited to the task of identifying hidden biomarkers from thousands of quantified proteins and has been used successfully to address problems as the prediction of genes associated with autosomal dominant disorders [11]. In this work, we first established and optimized a label-free-based quantitative proteomics protocol for renal FFPE biopsies and use this protocol to analyze a set of 15 FFPE biopsy samples including 5 TCMR, 5 BKPyVN and 5 STA. By combining the data collected from this label-free-based and the previous TMT-based proteomics experiments, a protein panel containing TCMR-specific biomarker was obtained. With different ML algorithms tested, an optimized ML-assisted model for precisely predicting of TCMR using kidney FFPE biopsies from renal allograft injury patients and normal controls was generated. We evaluated the performance of this prediction model using receiver operating characteristic (ROC) analyses to calculate its sensitivity and specificity using both of proteomics dataset and published microarray datasets deposited in the Gene Expression Omnibus website. Therefore, in this work, we made an effort to study FFPE biopsies from renal transplants using label-free-based quantitative proteomics profiling and ML to diagnose different kidney transplant injuries. Instead of using single biomarker for disease diagnosis, we attempted to use multi-biomarker panels to discriminate among biopsies belonging to different disease categories.

## 2. Materials and methods

### 2.1. Patients and sample collection

This study was approved by the University of Pittsburgh IRB (protocol 10110393). All patients received thymoglobulin induction with a rapid 7-day corticosteroid taper. Dual-maintenance immunosuppressive therapy consisted of mycophenolate mofetil and tacrolimus. Case selection was done from biopsies examined during routine clinical care over a 2-year period before initiation of this study. The principal author of this manuscript (P.R.) conducts a weekly biopsy conference that allows clinically validated diagnoses to be assigned to all renal allograft biopsies performed at the University of Pittsburgh. Five biopsies each were selected representing STA and TCMR. Biopsy designated as normal were protocol biopsies from stable patients. The core needle biopsy specimens (18 gauge) were fixed in formalin immediately and paraffin embedded within 24 hours.

### 2.2 Deparaffinization and protein extraction

The sample preparation to the FFPE biopsies was performed according to the methods described in previous studies with minor modifications. The biopsy tissue embedded in the paraffin blocks was extracted manually with a sharp scalpel, followed by cut into 1 mm pieces and placed in Protein LoBind Eppendorf tubes (Eppendorf, Hauppauge, NY, USA). The samples were then deparaffinized by incubating with xylene (Fisher Scientific, Pittsburgh, PA, USA) for 5 mins thrice and rehydrated with 100% ethanol for 3 mins thrice. After that, all samples were dissolved in an extraction buffer of 2% SDS dissolved with 20 mM Tris (pH8.0). Tissue was then mechanically disrupted by suction into a 3 mL syringe attached to an 18 gauge 1 ½ inch needle, followed by expulsion through a 23-gauge ½ inch needle into a conical tube on ice. The sample was then subjected to a focused ultrasonication step (work 4s, suspend 6s, total time 2min) with Model 120 Sonic Dismembrator (Fisher Scientific, Pittsburgh, PA, USA). The syringe disruption steps and the focused ultrasonication step were repeated alternately for a total of five times. The disrupted samples were incubated at 98 °C for 180 min, and supernatants collected by centrifugation at 10,000 × g for 10 min at 4 °C. With BCA assay measurement in triplicate, a 5-10 mm long needle core of kidney could yield 56.0-376.9 μg total protein. Unless otherwise noted, all other chemicals in this study were purchased from Sigma (St. Louis, MO, USA).

### 2.3 In-gel digestion

For each FFPE sample, 10 μg of protein were denatured and reduced with loading buffer containing 10 mM DTT at 37 °C for 1 h, alkylated with 25 mM iodoacetamide at room temperature for 30 min in the dark. Upon separation by SDS-PAGE and staining with Coomassie Blue (Roth), protein bands were excised from gels and subjected directly to tryptic digest. Tryptic digest was performed according to standard procedures with minor modifications [12]. Briefly, the gel pieces were sliced into 2 mm×2 mm gel pieces, destained with 50% acetonitrile (ACN, Merck) in 50 mM NH_4_HCO_3_ for three times and then washed with pure water for three times. Subsequently gel pieces were treated with pure ACN and rehydration with 50 mM NH_4_HCO_3_ buffer containing 2% ACN. Finally, the gel pieces were crushed and subjected to 50 mM NH_4_HCO_3_ buffer containing 10 ng/μl trypsin (Promega, Mannheim, Germany) with incubation over night at 37°C. Peptides were extracted with 80% ACN containing 0.1% formic acid and dried in a vacuum centrifuge. Then, the peptides were purified with stage-tip protocol [13] and dried in a vacuum centrifuge.

### 2.4 Liquid Chromatography with tandem mass spectrometry (LC-MS/MS)

Peptide separation was performed on a C18 capillary column (10.5 cm, 3 p,m, 120 Å) from New Objective (Woburn, MO, USA) under acidic conditions. The two eluent buffers were H_2_O with 2% ACN and 0.1% FA (A), and ACN with 2% H_2_O and 0.1% FA (B), and both were at pH 3. The gradient of the mobile phase was set as follows: 2%-35% B in 44 min, 35%-98% B in 1 min and maintained at 80% B for 3 min. The flow rate was 350 nL/min.

LC-MS/MS data was collected using an LTQ Orbitrap Velos mass spectrometer equipped with an ESI probe Ion Max Source with a microspray kit. The system was controlled by Xcalibur software version 1.4.0 from Thermo Fisher (Waltham, MA, USA) in the data-dependent acquisition mode. The capillary temperature was held at 320 °C, and the mass spectrometer was operated in positive ion mode. Full MS scans were acquired in the Orbitrap analyzer over the *m/z* 350-1,600 range with a resolution of 15,000 and the AGC target was 1e^6^. The 20 most intense ions were fragmented, and tandem mass spectra were acquired in the ion trap mass analyzer with. The dynamic exclusion time was set to 30 s, and the maximum allowed ion accumulation times were 60 ms for MS scans.

### 2.5 Data analysis

Raw data files were processed using Proteome Discoverer platform (Thermo Scientific, version 1.4) with SEQUEST as the search algorithms. MS/MS spectra were matched with a Uniprot *Homo sapiens* databases, using the following parameters: full trypsin digest with maximum 2 missed cleavages, static modification carbamidomethylation of cysteine (+57.021 Da), phosphorylation of serine, threonine and tyrosine as well as dynamic modification oxidation of methionine (+15.995 Da). Precursor mass tolerance was 10 ppm and product ions fragment ion tolerance were 0.8 Da. Peptide spectral matches were validated using percolator based on q-values at a 1% false discovery rate (FDR).

### 2.6 Machine learning and validation

Three machine learning predictive models were used: linear discriminant analysis (LDA), support vector machines (SVM), and random forest (RF). LDA uses Gaussian assumptions and Bayes theorem to estimate the posterior probability of being classified as TCMR for each testing sample [14]. Those with posterior probabilities greater than or equal to a specific cutoff are classified as TCMR. LDA was implemented by the “lda” function in the R package “MASS.” The second method SVM separates the STA and TCMR samples by finding a higher-dimension hyperplane that maximizes the margin, which is the minimum distance of the objects to the hyperplane [15]. SVM was implemented by the “svm” function in the R package “e1071.” RF classifies the samples by a majority vote of random trees using the classification and regression tree algorithm. The trees are constructed by bootstrapping of samples and subsampling of features [16]. This method was implemented using “randomForest” function in the R package “randomForest.” To evaluate the prediction performance of the protein signatures panel to distinguish TCMR from STA, we performed a leave-one-out cross-validation [17] and employed the above mentioned three learning algorithms (i.e. LDA, SVM and RF) respectively. Differential expression (DE) analysis to the training set with all protein features was performed using an empirical Bayes method by R package LIMMA. Protein features then were ranked based on their Benjamini-Hochberg (BH) adjusted p values. The subset of the features N ranged from 2 to 150 and the top N genes with smallest BH adjusted p values were selected to construct the model. Performance was evaluated by different perspectives including sensitivity, specificity and accuracy. The model was further validated on another independent 5 TCMR and 5 STA biopsies.

## 3. Results

### 3.1 Development of a Label-Free-Based Quantitative Proteomics Protocol for Kidney FFPE Biopsies

In our previous work, we reported a quantitative proteomic platform which was developed for molecular profiling of FFPE specimens [10]. The platform is consisted of a loss-less sample preparation method, a TMT10-plex-based quantitative proteomic workflow, and a systematic statistical analysis pipeline (**Figure 1A**). Quantitative comparison of the proteomes of a set of FFPE samples, including two renal allograft rejection diseases TCMR and BKPyVN, demonstrated that this TMT-based quantitative proteomics platform has excellent performance in differentiating various causes of renal allograft injury. However, the TMT-based platform may not be suitable for clinical practice considering the expensive labeling reagents and the tedious experimental procedures. In this present work, we developed and optimized a more clinic-friendly proteomics profiling protocol for renal FFPE biopsies with a label-free-based quantitative proteomics strategy. In this protocol, instead of labeling the tryptic peptides with TMT isobaric reagents followed by fractionation, the tryptic digests of FFPE specimens were injected directly to a LC column for LC-MS/MS analysis (**Figure 1B**). The raw LC-MS/MS data was subjected to quantitative analysis for peptides and proteins using the Proteome Discoverer software package. These identified and quantified proteins were then subjected to the systematic statistical analysis using bioinformatics tool of R package LIMMA to obtain differential expressed (DE) proteins (**Figure 1C**) before building a predictive model (**Figure 1D**). Using this protocol, we analyzed 15 additional FFPE biopsy samples including 5 TCMR, 5 BKPyVN and 5 STA. About 750–1250 proteins were identified and quantified with high confidence in each individual sample (**Supplementary Table S1-3**) using a 45 min LC gradient.

**Figure 1.**
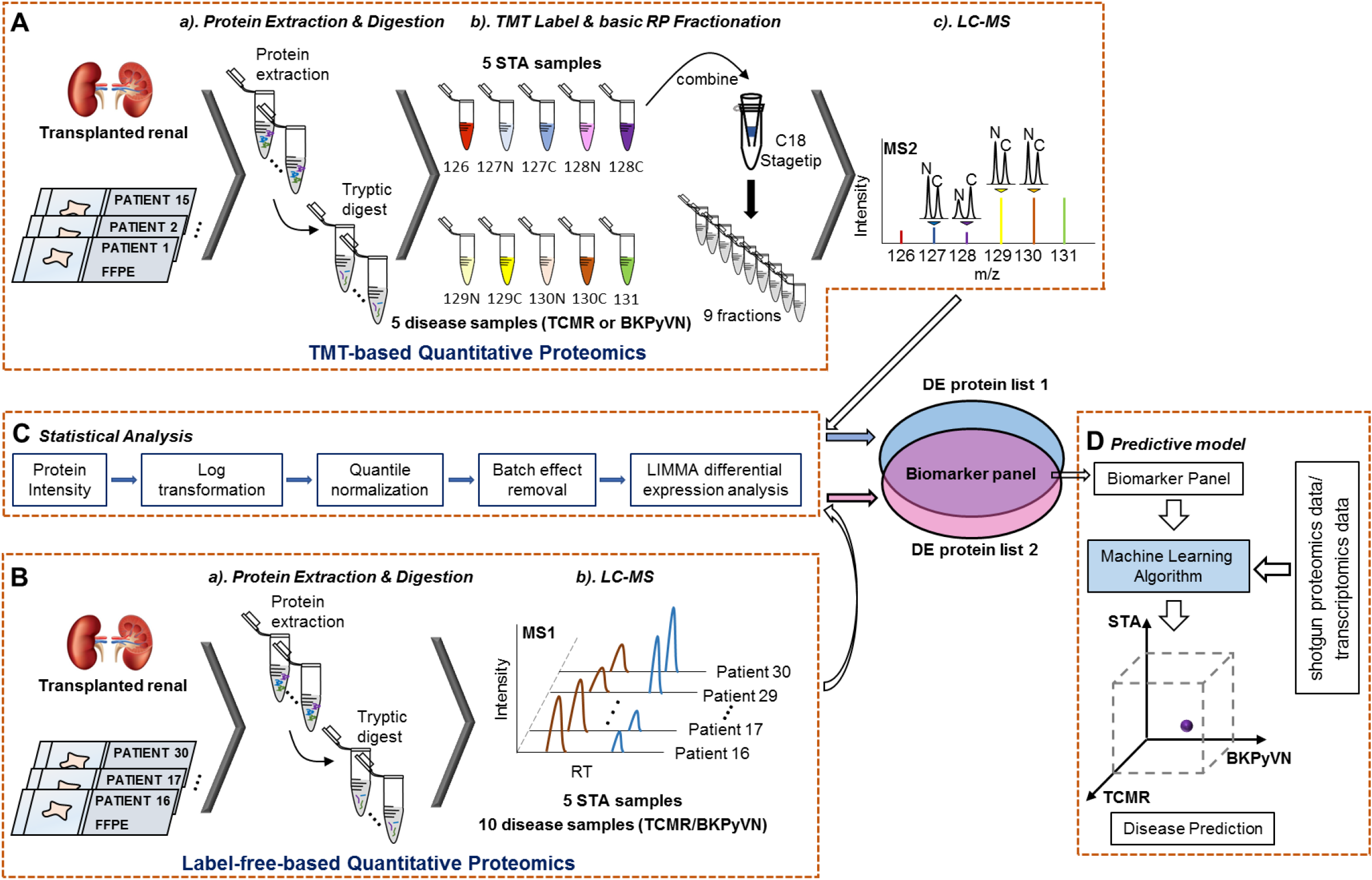
A flow chart for showing the procedures to diagnose of TCMR by FFPE biopsy-based proteomics and machine learning. **(A)** Experimental procedures for TMT-based quantitative proteomics. The proteins were extracted from 5 TCMR, 5 BKPyVN, and 5 STA biopsies, the digested peptides were labeled with TMT10-plex-reagents and separated by basic reverse phase C18 material. The fractionated peptides were subjected to LC-MS/MS analysis; **(B)** Experimental procedures for label-free-based quantitative proteomics. The proteins were extracted from another 5 TCMR, 5 BKPyVN, and 5 STA biopsies, the digested peptides were directly subjected to LC-MS/MS analysis; **(C)** The proteins were subjected to the systematic statistical analysis consisted of log transformation, quantile normalization, and LIMMA analysis to obtain differential expressed proteins; and **(D)** The machine learning algorithm was established based on the training data, and validated with testing data.

### 3.2 Label-Free-Based Quantitative Proteomics Analysis Distinguishes TCMR from STA and BKPyVN biopsies

Label-free-based proteomics is usually suffered from low repeatability. To remedy this defect in the label-free-based quantitative proteomics analysis of FFPE biopsies, we performed log transformation, quantile normalization, and batch effect removal before quantification analysis. As shown in **Figure 2A**, after data processing, a high Pearson’s correlation coefficient between the replicate experiments was achieved using our label-free-based protocol, demonstrating a good reproducibility in analyzing FFPE biopsy specimens.

**Figure 2.**
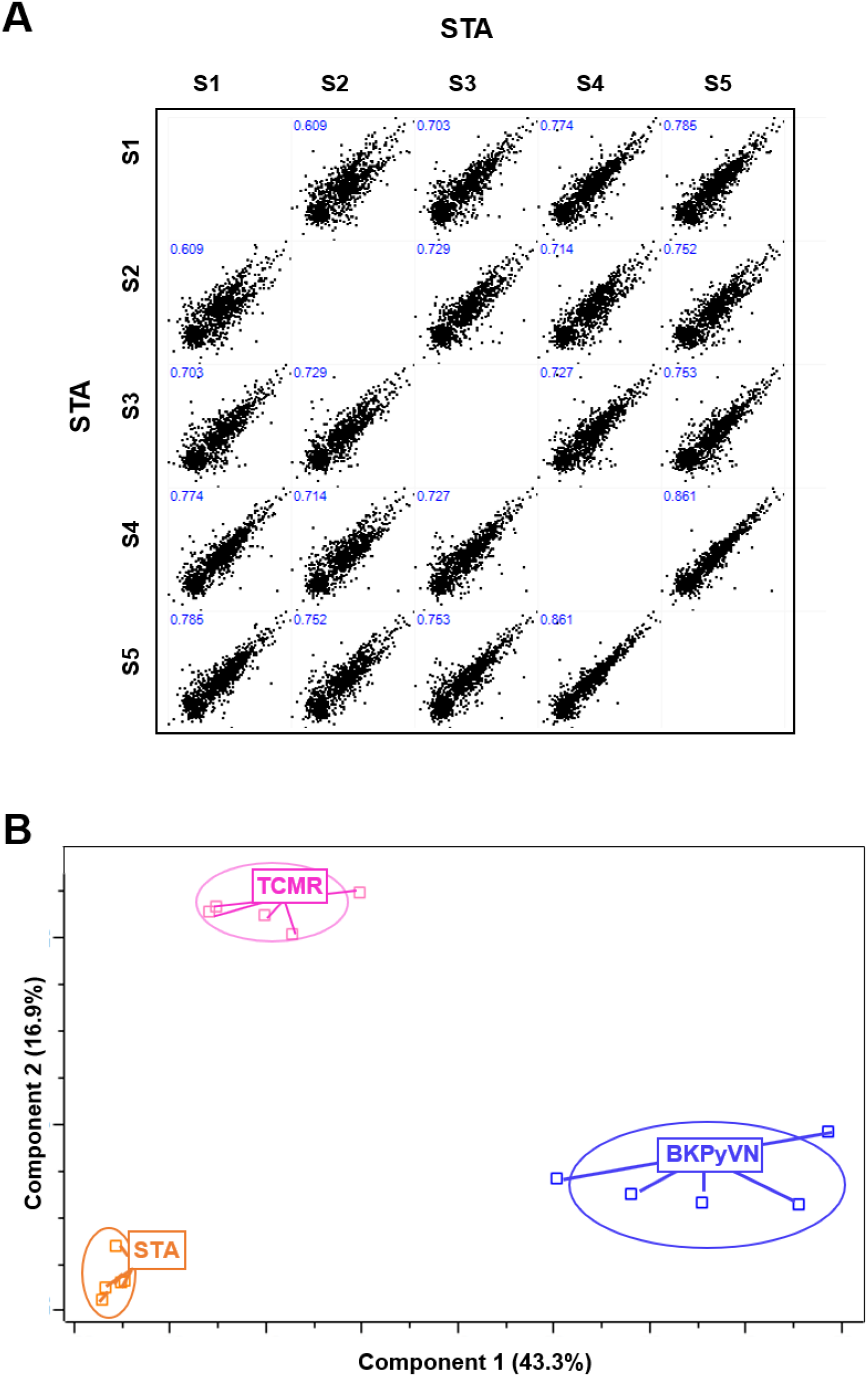
Quantitative proteomic profiling of FFPE biopsies segregates different allograft injuries. **(A)** Repeatability of label-free quantitative analysis. Correlations among 5 STA samples were shown. The correlation coefficient showed in the figure represents the statistical relationship between every two STA samples. The higher the number is, the higher repeatability between two samples is; **(B)** A PCA plot demonstrated that the quantified FFPE biopsy proteins were able to segregate STA, TCMR and BKPyVN samples. The PC1 axis is the first principal direction along which the samples show the largest variation. The PC2 axis is the second most important direction and it is orthogonal to the PC1 axis.

To test whether the label-free-based quantitative proteomics analysis could distinguish TCMR from other causes of kidney injuries, principal component analysis (PCA) was performed to the label-free-based proteomic profiling data obtained from STA, BKPyVN and TCMR biopsies (**Supplementary Table S4**). As shown in the **Figure 2B**, the quantified FFPE proteins not only segregate TCMR biopsies from control STA specimens (TCMR vs STA), but also distinguish the two tested disease phenotypes from each other (TCMR vs BKPyVN).

### 3.3 Differential Expression (DE) Analysis Reveals Potential Biomarkers for TCMR

To identify proteins in FFPE specimens that can serve as biomarkers to distinguish TCMR from other allograft injuries, DE analysis was performed using an empirical Bayes method implemented in R package LIMMA [18]. DE proteins were selected using two criteria: 1) their expression levels in TCMR biopsies significantly changed (i.e. the Benjamin–Hochberg procedure adjusted p value < 0.05) in comparison with STA samples at 1% FDR; 2) fold changes of protein expression levels between TC MR and STA are >2 or <-2. Totally, 178 out of the 778 quantified proteins were identified as DE proteins for TCMR when comparing to STA (**Supplementary Table S5**), with the expression levels of 42 proteins upregulated and 136 downregulated. Similarly, LIMMA analysis revealed that a total of 450 DE proteins significantly dysregulated in BKPyVN in comparison to STA samples (**Supplementary Table S5**), with the expression levels of 257 proteins upregulated and 193 downregulated. In addition, significant changes in expression levels of 281 proteins from TCMR occurred in comparison with BKPyVN biopsies (**Supplementary Table S5**).

### 3.4 Identification of Protein Classifiers for TCMR Suitable for Label Free-Based Proteomics Approach

To identify a specific and reliable protein signature panel for FFPE biopsies from TCMR patients, the common DE proteins that were confidently quantified with same trend (increase or decrease) in both label-free- and TMT-based proteomics analyses (**Supplementary Table S6-8**) were extracted. In this work, the STA sample was used as negative control for the disease samples. As a result, 32, 23, and 179 proteins were identified as common DE proteins in both two quantitative proteomics methods for TCMR vs STA, TCMR vs BKPyVN, and BKPyVN vs STA, respectively (**Table 1 & 2, Supplementary Table S9**). As shown in the reference sections in Table 1 and 2, a number of these proteins were previously reported to be associated with TCMR or BKPyVN. The results of bioinformatics analysis of these 32, 23, and 179 common DE proteins by Ingenuity Pathways Analysis are summarized in **Supplementary Table S13-15**.

**Table 1.** 32 DE proteins that were confidently quantified with same trend (increase or decrease) in both label-free- and TMT-based proteomics analyses for TCMR vs STA.

| Uniprot | Protein Names | Gene Names | logFC<br>-label<br>free | logFC<br>-TMT | Ref |
| --- | --- | --- | --- | --- | --- |
| **TCMR vs STA** |  |  |  |  |  |
| A6ND91 | Putative L-aspartate dehydrogenase | ASPDH | -2.02 | -0.28 | [36] |
| O00764 | Pyridoxal kinase | PDXK | -3.06 | -0.37 | [37] |
| O43598 | 2'-deoxynucleoside 5'-phosphate N-hydrolase 1 | DNPH1 | -3.21 | -0.62 |  |
| P01034 | Cystatin-C | CST3 | 2.67 | 0.82 | [38] |
| P02042 | Hemoglobin subunit delta | HBD | -1.59 | -0.82 | [39] |
| P02790 | Hemopexin | HPX | -2.44 | -0.57 |  |
| P02792 | Ferritin light chain | FTL | 1.50 | 1.43 | [40] |
| P05937 | Calbindin | CALB1 | -3.74 | -0.97 | [41] |
| P07585 | Decorin | DCN | 2.16 | 1.15 | [42] |
| P08670 | Vimentin | VIM | 1.36 | 0.69 | [43] |
| P11177 | Pyruvate dehydrogenase E1 component subunit beta, mitochondrial | PDHB | -2.11 | -0.32 | [44] |
| P12109 | Collagen alpha-1(VI) chain | COL6A1 | 2.98 | 0.46 | [45] |
| P13796 | Plastin-2 | LCP1 | 2.30 | 0.68 | [46] |
| P14866 | Heterogeneous nuclear ribonucleoprotein L | HNRNPL | 1.69 | 0.21 | [47] |
| P25787 | Proteasome subunit alpha type-2 | PSMA2 | -2.75 | -0.30 | [48] |
| P30043 | Flavin reductase (NADPH) | BLVRB | -3.32 | -0.53 | [49] |
| P38919 | Eukaryotic initiation factor 4A-III | EIF4A3 | 2.43 | 0.29 |  |
| P49789 | Bis(5'-adenosyl)-triphosphatase | FHIT | -1.99 | -0.29 |  |
| P50053 | Ketohexokinase | KHK | -1.92 | -0.30 | [36] |
| P50454 | Serpin H1 | SERPINH1 | 2.21 | 0.48 | [36] |
| P51606 | N-acylglucosamine 2-epimerase | RENBP | -2.56 | -0.48 | [50] |
| Q07507 | Dermatopontin | DPT | 2.46 | 0.89 | [51] |
| Q14376 | UDP-glucose 4-epimerase | GALE | -2.07 | -0.27 | [52] |
| Q14894 | Ketimine reductase mu-crystallin | CRYM | -1.46 | -0.38 | [53] |
| Q15582 | Transforming growth factor-beta-induced protein ig-h3 | TGFBI | 2.37 | 0.42 | [54] |
| Q16795 | NADH dehydrogenase [ubiquinone] 1 alpha subcomplex subunit 9, mitochondrial | NDUFA9 | -2.06 | -0.35 | [55] |
| Q5R3I4 | Tetratricopeptide repeat protein 38 | TTC38 | -2.65 | -0.31 | [56] |
| Q86YB7 | Enoyl-CoA hydratase domain-containing protein 2, mitochondrial | ECHDC2 | -3.06 | -0.57 | [56] |
| Q93099 | Homogentisate 1,2-dioxygenase | HGD | -1.15 | -0.32 | [57] |
| Q96C23 | Aldose 1-epimerase | GALM | -1.33 | -0.60 |  |
| Q9BSH5 | Haloacid dehalogenase-like hydrolase domain-containing protein 3 | HDHD3 | -3.54 | -0.81 | [56] |
| Q9Y2S2 | Lambda-crystallin homolog | CRYL1 | -1.50 | -0.38 | [58] |

**Table 2.** 23 DE proteins that were confidently quantified with same trend (increase or decrease) in both label-free- and TMT-based proteomics analyses for BKPyVN vs TCMR.

| Uniprot | Protein Names | Gene Names | logFC<br>-label<br>free | logFC<br>-TMT | Ref |
| --- | --- | --- | --- | --- | --- |
| **BKPyVN vs TCMR** |  |  |  |  |  |
| O00571 | ATP-dependent RNA helicase DDX3X | DDX3X | 2.28 | 0.30 | [59] |
| O15145 | Actin-related protein 2/3 complex subunit 3 | ARPC3 | 2.41 | 0.48 | [60] |
| P04080 | Cystatin-B | CSTB | 2.49 | 0.23 | [44] |
| P05388 | 60S acidic ribosomal protein P0 | RPLP0 | 1.88 | 0.64 | [61] |
| P12429 | Annexin A3 | ANXA3 | 1.48 | 0.42 | [62] |
| P21397 | Amine oxidase [flavin-containing] A | MAOA | -2.09 | -0.40 | [63] |
| P30043 | Flavin reductase (NADPH) | BLVRB | 2.76 | 0.35 | [49] |
| P30740 | Leukocyte elastase inhibitor | SERPINB1 | 2.13 | 0.58 | [64] |
| P35914 | Hydroxymethylglutaryl-CoA lyase,<br>mitochondrial | HMGCL | -2.72 | -0.39 |  |
| P36578 | 60S ribosomal protein L4 | RPL4 | 1.93 | 0.27 | [65] |
| P39656 | Dolichyl-diphosphooligosaccharide--protein<br>glycosyltransferase 48 kDa subunit | DDOST | 1.62 | 0.23 |  |
| P47755 | F-actin-capping protein subunit alpha-2 | CAPZA2 | 3.66 | 0.28 |  |
| P62277 | 40S ribosomal protein S13 | RPS13 | 3.56 | 0.20 | [66] |
| P62913 | 60S ribosomal protein L11 | RPL11 | 4.17 | 0.32 | [67] |
| P84103 | Serine/arginine-rich splicing factor 3 | SRSF3 | 3.31 | 0.31 |  |
| Q02878 | 60S ribosomal protein L6 | RPL6 | 1.80 | 0.32 | [68] |
| Q07507 | Dermatopontin | DPT | -2.26 | -0.48 | [51] |
| Q14974 | Importin subunit beta-1 | KPNB1 | 2.73 | 0.30 | [69] |
| Q7KZF4 | Staphylococcal nuclease domain-containing<br>protein 1 | SND1 | 2.22 | 0.31 | [70] |
| Q92945 | Far upstream element-binding protein 2 | KHSRP | 1.83 | 0.19 | [71] |
| Q96NB2 | Sideroflexin-2 | SFXN2 | -3.63 | -0.51 |  |
| Q9UJ70 | N-acetyl-D-glucosamine kinase | NAGK | 1.98 | 0.42 | [72] |
| Q9Y6C2 | EMILIN-1 | EMILIN1 | 2.43 | 0.44 | [36] |

### 3.5 Comparison of Different Machine Learning Algorithms and for Construction of a Prediction Model for TCMR, BKPyVN and STA

Predictive modeling is a method of creating models that can identify the likelihood of disease. Within the modeling, machine learning algorithms employ a variety of statistical, probabilistic and optimization methods to learn from known knowledge and to detect useful patterns from large datasets that relies on categorized training data [19]. In this work, to develop a prediction model that can distinguish TCMR, BKPyVN and STA, the 234 (32 + 23 + 179) DE proteins commonly quantified from both TMT- and label-free-based quantitative proteome analyses (**Table 1 & 2, Supplementary Table S9**) were used as the classifiers. The detailed procedures to construct the predictive model are outlined in **Figure 3**. Three different machine learning algorithms, i.e. linear discriminant analysis (LDA), support vector machine (SVM) and random forests (RF), were respectively applied to the protein panel and the performance of these machine learning algorithms was compared by leave-one-out cross-validation. In each cycle of cross-validation, one sample was held as the evaluation set and the other fourteen samples as training set. As shown in **Figure 4**, disease and normal phenotypes could be accurately and obviously distinguished using the three prediction models we developed, with 100%, 100% and 93.3% accuracy achieved in cross-validation for SVM, RF and LDA, respectively. The receiver operating characteristic (ROC) curve, which has been widely used in clinical epidemiology, was also performed to quantify how accurately our prediction model for discriminating between "diseased" and "non-diseased" states [20]. For all three algorithms, the area under the curve (AUC) of 1 for the injury subtype provides 100% specificity and 100% sensitivity between each two disease types (**Supplementary Figure S1**) [21].

**Figure 3.**
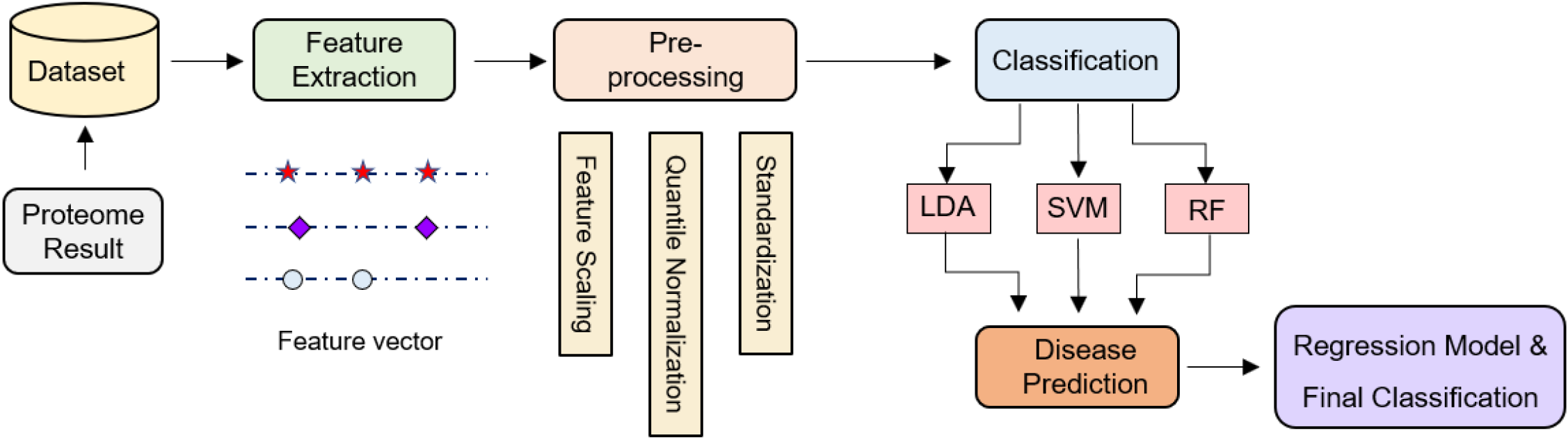
Development of the machine learning derived disease prediction model. Feature/Attribute selection process selects the critical features for the prediction of renal allograft rejection disease. After feature selection, preprocessing involved to remove the outlier and make dataset normalized. Various classification techniques were applied to preprocessed data. Finally, model evaluation is performed based on different measures.

**Figure 4.**
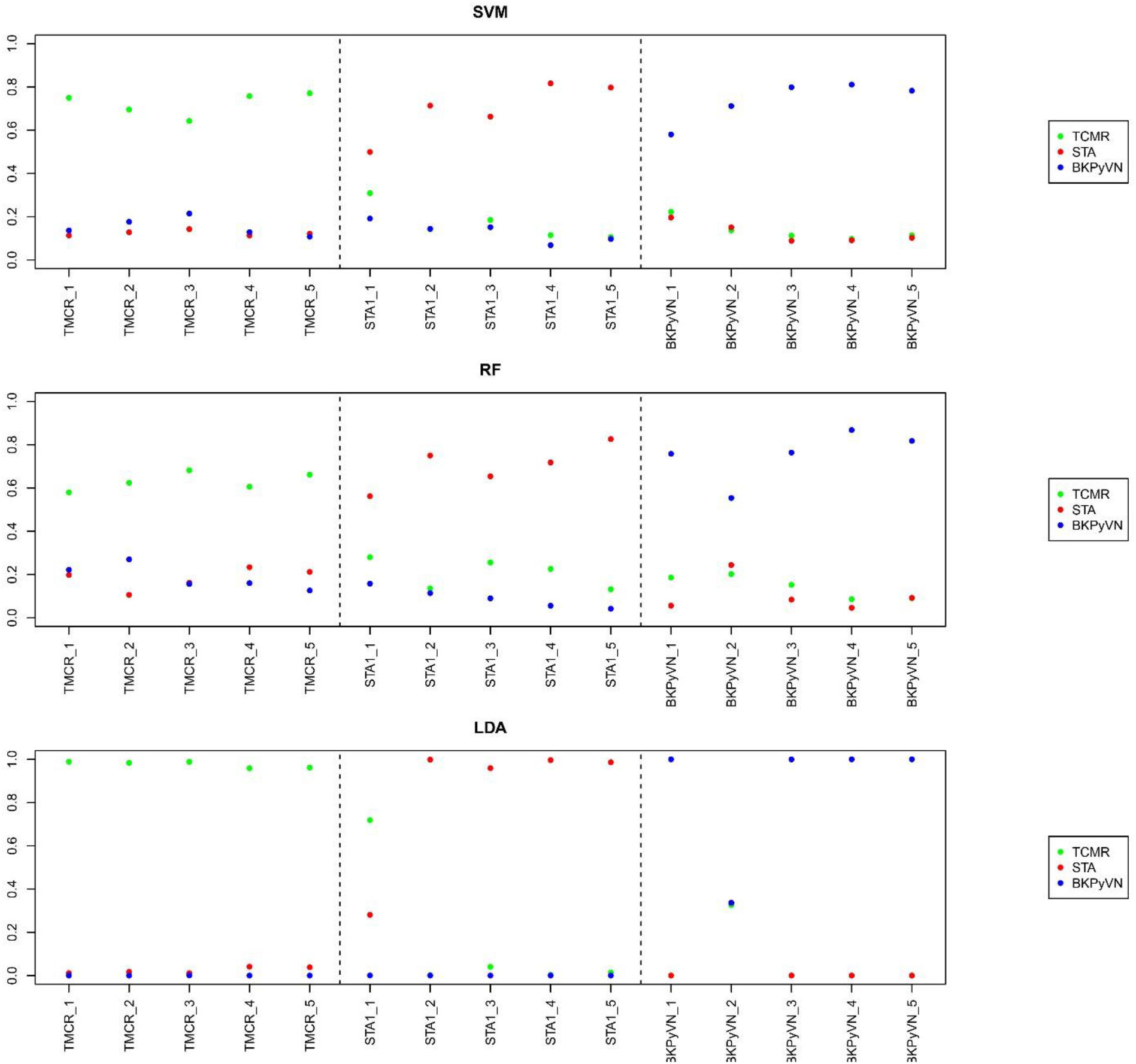
Diagnostic ability of the three different predictive models applied to disease and normal phenotypes. The probability calculated for the renal allograft injuries using biomarker panel with the three different prediction models.

### 3.6 Validation of the Prediction Model Using Published Transcriptome Datasets

To further ensure its feasibility, the transcriptome data was used to test the performance of our predictive model for TCMR from STA. The classifiers using the 32 DE proteins commonly quantified from both TMT- and label-free-based quantitative proteome analyses (**Table 1**) were applied to two microarray–based datasets (GSE48581[22] and GSE36059 [23]) posted on the Gene Expression Omnibus website. Applying the aforementioned three predictive models to GSE36059 achieves 26/35=74% (SVM), 27/35=77% (RF) and 25/35=71.4% (LDA) in sensitivity as well as 157/281=55.9% (SVM), 176/281=62.6% (RF) and 172/281=61.2% (LDA) in specificity, respectively. Meanwhile, when applied to GSE48581, the sensitivities of the three models are 25/32=78% (SVM), 23/32=71.8 (RF) and 22/32=68.8% (LDA) as well as the specificities are 135/222=60.8% (SVM), 142/222=64.0% (RF) and 135/222=60.8% (LDA), respectively. Furthermore, the integration of the three predictive models and the classifier containing 234 (32+179+23) DE proteins commonly obtained from both TMT- and label-free-based quantitative proteome analysis were performed to GSE72925 [24] to distinguish TCMR, BKPyVN and STA. In this dataset, a total of 99 testing samples (66 STA + 5 BKPyVN + 26 TCMR) were analyzed. As shown in **Supplementary Table S16**, in comparison with the SVM (40%) and LDA (29%), the RF-based model achieved the highest accuracy as 47%.

## 4. Discussion

With current immunosuppressive therapy, acute rejection develops in about 10%-12% of transplant patients [25]. TCMR, which is a cognate recognition-based process that creates local inflammation and epithelial dedifferentiation, stereotyped nephron responses, and tubulitis, will cause irreversible nephron loss if untreated [2]. Cherukuri et. al. [26] reported that patients with clinical TCMR have significantly worse graft outcomes (allograft chronicity at 1 year and impending graft loss) in comparison to those without TCMR. However, due to the acknowledged limitations of conventional diagnostic systems, which are based on histologic lesions interpreted by empirically derived guidelines moderated by Banff consensus [3], there’s an urgent need to develop precision diagnostics to TCMR.

Disease prediction modeled by machine learning is on the rise due to their potential for advanced predictive analytics, which is creating many new opportunities for healthcare. Briefly, the supervised model of learning aimed to predict the value of a variable called output variable from a set of variables called input variable (**Figure 3**). In this work, the feature vector, as the basic building blocks of datasets, was composed of protein name and the corresponded intensity in three biopsies. The set of input and output variables were used as training and testing data. Training data is the known data, whereas testing data is the unknown data to be predicted. Firstly, we need to determine the input variable source. FFPE of tissues preserves the morphology and cellular details of tissue samples. Thus it has become the standard preservation procedure for diagnostic surgical pathology [27]. The commonly used approach with FFPE tissue for diagnosis, the transcriptome analysis, is ambiguous because the DNA from FFPE biopsies is often highly cross-linked, degraded and fragmented [5]. Meanwhile, it has been reported that there is no significant difference between macromolecules, especially proteins, extracted from FFPE samples stored over 10 years in comparison with the current year blocks [27, 28], which is beneficial for us to take advantage of this readily available resource. Thanks to the dramatically improvement in LC separation and MS instruments [29, 30], proteomics research becomes a rapidly growing field in holding the promise of discovery of biomarkers of acute rejection and elucidation of pathophysiologic mechanism of rejection [31].

In our previous work, a TMT-based quantitative proteomic platform was successfully developed for molecular profiling of FFPE specimens [10]. Comparing to the label-free-based proteomic strategy, the TMT-based approach provides a more accurate way to quantify and compare proteomes in biological samples. By chemical labeling (or tagging) the peptides from different samples with specific but different isobaric mass tags, peptides prepared from multiple samples can be pooled for a single analysis since the mass spectrometry can differentiate these peptides due to the differences in the mass tags [32]. For example, the TMT10-plex kit we used in our previous report for FFPE biopsies contains a set of ten isobaric mass tags, allowing the analysis of 10 samples in one experiment to improve the quantitative accuracy. However, there are limitations when introducing the TMT-based proteomic approach to clinical practice. First, only a limited number of samples can be compared in one TMT-based experiment. Currently, the maximum number of samples can be used in a TMT-based experiment is 16 by using the TMTpro-16plex kit. Second, these TMT-labeling reagents are expensive. Third, the TMT-based labeling procedures are labor-intensive and the quantitative accuracy can be sacrificed by low labeling efficiency if the experiments are not performed in optimized conditions. Therefore, in this current work, we further developed a label-free-based quantitative proteomic analysis protocol for FFPE biopsies, as a more clinic-friendly tool than the TMT-based method, considering the advantages of simplified experimental procedures and the possibility of performing comparative quantification across many samples. In addition, once the label-free proteomics-based clinical test is developed and validated, the cost for reagents can be as low as a few dollars per test. Therefore, we estimated if a platform integrating of a label-free-based quantitative proteomics technology and machine learning algorithms could provide a proteomic profiling “fingerprints” with a panel of protein classifiers. With the information of protein classifiers, we could establish a prediction model that can accurately differentiate the TCMR biopsies from other kidney transplant injuries.

The label-free-based quantitative proteomic strategy was performed to the 15 FFPE biopsy samples including 5 TCMR, 5 BKPyVN and 5 STA. The high Pearson’s correlation coefficient between the replicate experiments demonstrated that a good reproducibility can be achieved using this method. The PCA clustering result revealed that the label free-based proteomic profiling data in combination with strict bioinformatics analysis of FFPE specimens is capable of distinguishing among different allograft injuries. By using bioinformatics tool of R package LIMMA, label-free-based DE protein lists among TCMR, STA and BKPyVN samples were obtained. To obtain a panel of protein classifiers/biomarkers to diagnose TCMR with higher accuracy, we chosen the DE proteins confidently quantified with same expression level trend (increase or decrease) in label-free-based- and TMT-based proteomics analyses. As a result, 32, 23, and 179 proteins were identified as common DE proteins for TCMR vs STA, TCMR vs BKPyVN, and BKPyVN vs STA, respectively. The protein intensity data (the summarized intensities of all identified peptides for each protein) in the FFPE biopsies of STA, TCMR and BKPyVN obtained from the label-free-based experiments was used as the classifiers for machine learning prediction model.

Among the protein classifiers/biomarkers chosen in this study, the 32 common DE proteins between TCMR and STA include a number of proteins associated with renal inflammation, damage, tubule injury, nephritis, and nephrosis, such as cystatin C (increased), decorin (increased), hemopexin (decreased), and crystallin mu (decreased). Cystatin C, an extracellular space protein, has been used as a biomarker for diagnosis of kidney function (glomerular filtration rate, GFR) (the identifier in the ClinicalTrials.gov database (https://clinicaltrials.gov/) as NCT00300066) and for prognosis of ischemic stroke (NCT00479518). This protein was also been used as a biomarker for measuring the efficacy of valsartan in treatment of hypertension for patients with renal dysfunction (NCT00140790). In addition to cystatin C, there are several other proteins among the 32 common DE proteins for TCMR vs STA that have been used as or are potential biomarkers for clinical diagnosis. These proteins are vimentin, lymphocyte cytosolic protein 1, homogentisate 1,2-dioxygenase, and ferritin light chain. Ingenuity Pathways Analysis (IPA) of these 32 common DE proteins revealed two cellular protein networks. One network is associated with Cell Morphology, Cellular Assembly and Organization, Cellular Function and Maintenance (**Figure 5A and Supplementary Table S13**) and the other one is associated with Cell Cycle, Gene Expression, Cell-To-Cell Signaling and Interaction (**Figure 5B and Supplementary Table S13**). Ingenuity Pathways Analysis of the 23 common DE proteins for TCMR vs BKPyVN suggests that these proteins are involved protein synthesis, RNA damage and repair, cell death and survival (**Supplementary Table S14**). Among these 23 proteins, cystatin B, annexin A3, and DEAD-box helicase 3 X-linked have been used as biomarkers for cancer in clinical diagnosis. Two cellular networks were enriched for these DE proteins between TCMR and BKPyVN. One network is associated with Protein Synthesis, RNA Damage and Repair, and Cancer and other one is associated with Cell Cycle, Energy Production, and Molecular Transport. The bioinformatics findings provide new insights into the underlying mechanisms for the development of these kidney allograft injuries.

**Figure 5.**
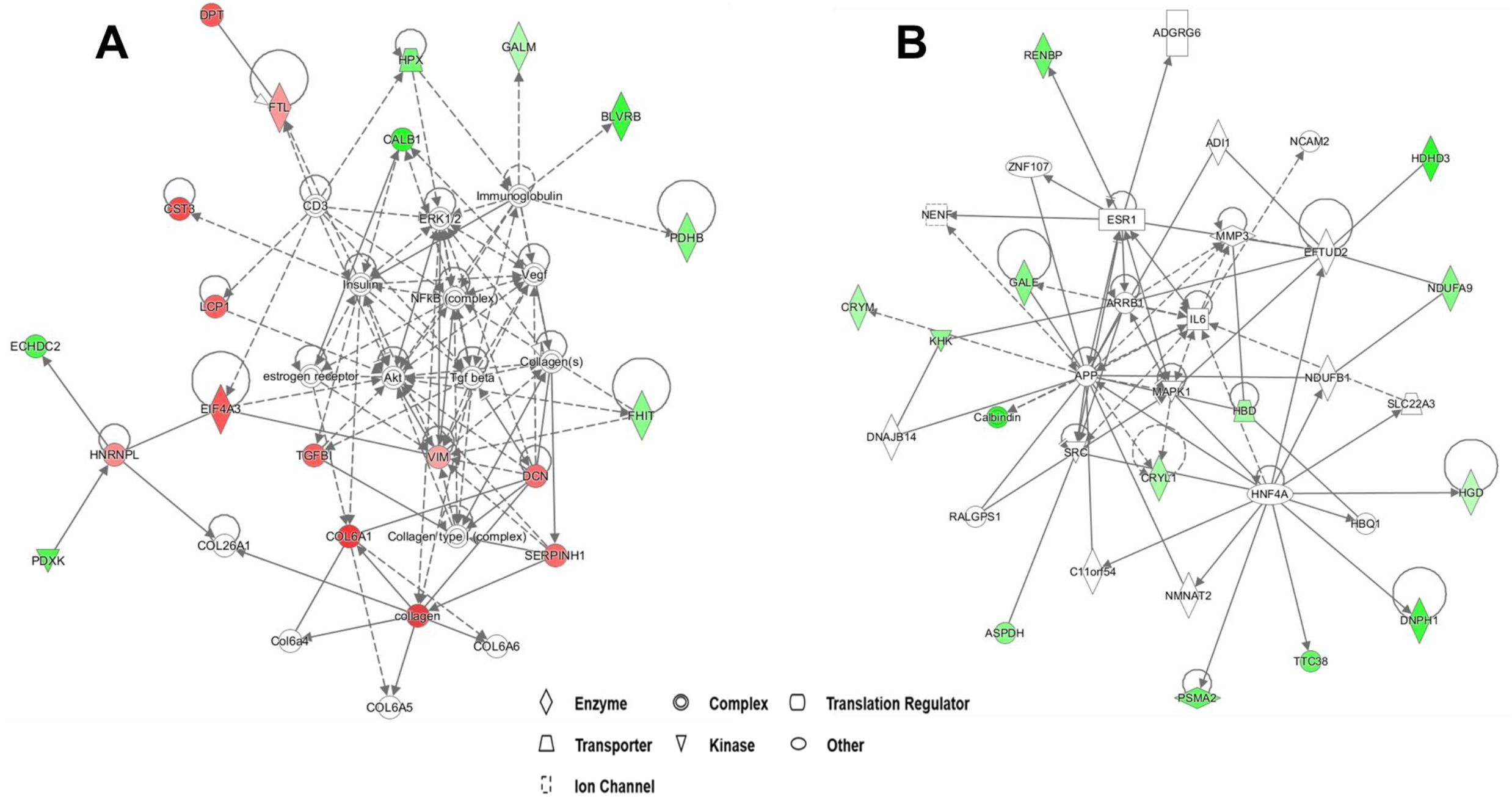
Ingenuity Pathways Analysis of the common DE proteins for TCMR vs STA reveals cellular networks associated with TCMR. **(A)** Network of Cell Morphology, Cellular Assembly and Organization, Cellular Function and Maintenance. **(B)** Network of Cell Cycle, Gene Expression, Cell-To-Cell Signaling and Interaction.

As the core component of developed prediction model, the selection of an optimal machine learning algorithm is prerequisite. Logistic regression (LOR), Decision tree (DT), Random forest (RF), k-Nearest Neighbors (k-NN), Support vector machine (SVM), Naive Bayes (NB) and Artificial neural network (ANN) are among the most commonly used machine learning techniques [33–35]. In this study, three machine learning algorithms, LDA, SVM, and RF, were applied to quantitative proteomics data collected from renal FFPE biopsies. To test the three algorithms, the 234 DE proteins commonly quantified from both TMT- and label-free-based quantitative proteome analysis was performed as training data. With leave-one-out cross-validation, all three algorithms were found to achieve excellent predictive performance for rejection with 100% sensitivity and specificity, demonstrating that a high diagnostic potential of using our prediction model to discriminate the true state of subjects. In addition, the model was also applied to predict the transcriptome data with high sensitivity and specificity, with the RF-based model achieved the highest accuracy in prediction.

Although our study sample size was small, there is certainly no simple rule of thumb to determine the necessary sample size for the omics study to find novel biomarkers. However, rejection is a heterogeneous process. Although we applied stringent histopathologic criteria to define acute TCMR, a larger sample size might be necessary to cover the broad spectrum of TCMR.

In conclusion, we successfully developed an integrative pipeline by integrating label-free-based quantitative proteomic analysis and machine learning derived prediction model for TCMR diagnosis. Subsequent validation of the proteomic discoveries by shotgun analysis with blindly test biopsies confirmed that the developed model could serve as a potential diagnostic tool for acute TCMR. To the best of our knowledge, this is the first time to provide a proteomics-based diagnostic method with FFPE biopsies for distinguishing TCMR from STA samples. To further demonstrate the clinical effectiveness of the obtained biomarker panel, appropriately powered clinical trials with a sufficient number of TCMR and control patients, as well as a sufficient study period are deemed necessary in the near future.

## Supporting Information

Supporting Information is included and available from the author.

## Data Availability

All data referred to in the manuscript will be available upon request.

## Acknowledgements

This publication was also made possible by seed funding support to K.X. from the Department of Pharmacology and Chemical Biology, the University of Pittsburgh and Vascular Medicine Institute, the Hemophilia Center of Western Pennsylvania, and the Institute for Transfusion Medicine.

## Abbreviations

FFPE: formalin-fixed and paraffin embedded
STA: kidney tissue with stable function
TCMR: T-cell mediated rejection
BKPyVN: polyomavirus BK nephropathy
DE: differentially expression

